# Hospital exposure to body fluids and vaccination status of caregivers: a cross-sectional study in public hospitals of Yaoundé-Cameroon

**DOI:** 10.1101/2025.09.10.25335527

**Authors:** Rick Tchamani, Armel Evouna Mbarga, Claude Axel Minkandi, Guy-Roger Ebanda, Fabrice Zobel Lekeumo Cheuyem

## Abstract

**Background:** In many Sub-Saharan African countries, structural health system challenges lead to a heavy reliance on informal caregivers to provide basic patient care in hospitals. These caregivers are frequently exposed to blood and body fluids (BBF), placing them at high risk of infections like hepatitis B, COVID-19, and cholera. This study aimed to assess the prevalence and determinants of BBF exposure and vaccination coverage among informal caregivers in public hospitals in Yaoundé, Cameroon.

**Methods:** A cross-sectional study was conducted between April and July 2025 at two public reference hospitals in Yaoundé. A total of 327 caregivers, aged ≥21 and caring for a hospitalized patient for ≥48 hours, were enrolled using a non-probabilistic convenience sampling technique. Data on sociodemographics, hospital experiences, BBF exposure, and vaccination status were collected via face-to-face interviews using a pre-tested questionnaire. Multivariate binary logistic regression was used to identify factors associated with full vaccination and exposure to BBF.

**Results:** A high proportion (57.49%) reported exposure to BBF, most commonly to urine (87%), expectoration (38%), and vomit (21%). Vaccination coverage was critically low: only 10.40% were fully vaccinated against hepatitis B, 6.12% against COVID-19, and 3.98% against cholera. Furthermore, 67.59% had not received any of the three vaccines. Multivariate analysis revealed female caregivers were significantly less likely to be fully vaccinated against COVID-19 (aOR=6.27), hepatitis B (aOR=2.98), and cholera (aOR=3.36) compared to males. Higher education level was associated with lower COVID-19 vaccine uptake (aOR=29.7), and unemployed caregivers were less likely to be vaccinated against hepatitis B than public sector workers (aOR=6.43). Caregivers who performed tasks had seventeen-fold increased odds of BBF exposure (aOR=17.1). Those with prior caregiving experience were three times more likely to be exposed (aOR=2.98). Married status and reporting difficulty sleeping were also significantly associated with higher exposure risk to BBF.

**Conclusion:** Informal caregivers in Cameroonian hospitals face high exposure to blood and body fluids, coupled with low vaccination coverage against key infectious diseases. These results underscore an urgent need for public health policies to recognize and protect them, including integrating them into infection prevention programs and implementing targeted vaccination campaigns within healthcare facilities.

## Introduction

Health systems in most Sub-Saharan African countries face major structural challenges, including a shortage of qualified personnel, insufficient budget allocations, and a deficit in leadership and management [1,2]. These constraints directly affect the availability and quality of medical equipment and facilities in hospitals.

As a result of a persistent shortage of qualified healthcare staff to meet the demand for care, a significant portion of basic care is provided by patients’ families and close acquaintances within healthcare settings [3,4]. These informal caregivers, who are compelled to actively participate in patient care, take on essential tasks such as bathing, feeding, and administering medication [5].

Consequently, due to their constant presence at the patient’s bedside in the hospital setting and the absence of adequate protective measures, informal caregivers are vulnerable to direct exposure to blood and other bodily fluids. They therefore run a high risk of contracting life-threatening infections, such as HIV, hepatitis B, hepatitis C, COVID-19 or other directly transmitted bacterial infections like cholera.

Hospital transmission can occur through splashes to the mucous membranes (eyes, nose, mouth) or broken skin, but also through percutaneous injuries such as needle sticks and cuts [6,7]. Their lack of knowledge about transmission mechanisms significantly exacerbates this risk [8].

This infectious peril exists in a context where Cameroon faces a significant burden of infectious diseases, compounding its public health challenges. In 2023, the national hepatitis B prevalence was 11.9% [9]. Cholera and COVID-19 outbreaks were particularly severe, resulting in 478 and 1,974 deaths, respectively, with high case fatality rates of 2.7% and 1.6% [10,11]. Despite this high burden, vaccine coverage remains inadequate [12–14], with public acceptance especially low for cholera and COVID-19 vaccines [15,16].

In addition to the direct infectious risk, caregivers face precarious living conditions within hospitals, where they often sleep on benches, in corridors, or even outdoors [17]. This overcrowding doubly increases their vulnerability by exposing them to pathologies such as malaria, the common cold, or flu-like syndromes.

Thus, in their show of solidarity, these caregivers find themselves plunged into an environment that not only jeopardizes their health but also risks making them ill in turn. This problem stems directly from the structural shortcomings of the Cameroonian health system. Indeed, it does not officially recognize the place of the informal caregiver in the hospital, and access to health services is primarily ensured through direct “out-of-pocket” payments by users. This results in multiple barriers to accessing care, translating into incomplete universal health coverage, an extremely low insurance rate, and insufficient household income to procure quality services [18].

Faced with this new hospital environment, which must take this population and their health into account, it is necessary to implement sound policies to protect informal caregivers within hospitals. The establishment of such programs requires an assessment of their exposure risks. However, data that describe the experience of this specific population within Cameroon’s health facilities are scarce. This cross-sectional study therefore aimed to provide a descriptive assessment and identify determinants of hospital exposure to blood and other body fluids among caregivers, as well as their vaccination status.

## Methods

### Study design & period

Between April and July 2025, we conducted a cross-sectional study for descriptive and analytical purposes at two public reference hospitals in Yaoundé: the Biyem-Assi District Hospital and the Military Hospital of Region No.1.

### Setting

The study was set in Yaoundé, the political capital of Cameroon, a city of 1.5 million people that hosts all the nation’s ethnic groups. Its demographic profile is notably young and has a gender imbalance skewed towards men, with the majority (70-80%) [19]. The selected study sites were two public reference hospitals: the Biyem-Assi District Hospital (a first-level referral center in the Yaoundé VI subdivision) and the Military Hospital of Region No. 1 (a third-level referral center in the Yaoundé III subdivision) [20]. In 2024, these two institutions had a combined capacity of 300 beds and managed approximately 73,000 consultations and 9,900 hospitalizations, providing over 35,000 days of inpatient care [21].

### Study participants & selection criteria

Eligibility required individuals to be at least 21 years old and a caregiver for a patient hospitalized in one of the selected facilities for ≥48 hours. All eligible caregivers who provided informed consent were included.

### Sampling method

The sample size was calculated using the single proportion formula, n=(Zα/2)2P(1−P)/d2, at a 95% confidence interval. In this formula, Zα/2=1.96, P=50% prevalence (since no similar study had been conducted previously in the study area), and d=5% marginal error. Using the formula, an estimated sample size of n=384 was obtained. The finite population correction was then applied to adjust the initial sample size (n) because the sampling frame was small (N≈1500 caregivers during the 3-month period) [22]. The adjusted sample size was calculated using the formula: Adjusted n=(n×N)/(n+(N−1)), where n is the initial sample size (384) and N is the total number of eligible caregivers in the sampling frame during the study period. The calculation is as follows: Adjusted n = (384×1500)/ (384+(1500−1)) = 576,000/1883 ≈ 306. Thus, the required sample size was adjusted down to 306. Caregivers were approached in their respective departments and asked to participate. A non-probabilistic convenience sampling technique was used to enroll consenting caregivers.

### Data collection tool and procedure

The study tool was a questionnaire designed to collect sociodemographic, hospital stay experience, hospital exposure to body fluids, and vaccination status. The questionnaire was pre-tested and then administered through face-to-face interview method with consenting caregivers.

### Variables

The dependent variables included exposure to body fluids during the hospital stay and full vaccination status against hepatitis B, COVID-19, and cholera. The independent variables encompassed sociodemographic characteristics (age, gender, education level, marital status, occupation, and income) and hospital experience (tasks performed during the hospital stay, experience trouble with a healthcare worker, prior experience as a caregiver, and perceived ease of sleeping during the hospital stay), as well as compliance with full vaccine uptake.

### Data processing and analysis

Data were entered, exported, recoded as necessary, and analyzed using R Statistics version 4.4.2 [23]. The respondent characteristics were presented as counts and frequencies. Simple and multiple binary logistic regressions were used to assess the strength of association between variables. The selection of predictors that best fit the model was done stepwise using the Akaike Information Criterion (AIC)[24]. The model with the lowest index was then selected. A *p*-value <0.05 was considered statistically significant.

### Ethical approval statement

This protocol was approved by the Human Health Research Ethical Review Committee for the Centre Region (CRERSH - Ce) and the ethical clearance: CE Nº 00379/CRERSH/2025 issued. Informed consent was obtained from participants prior to inclusion in the study. All methods were performed in accordance with declaration of Helsinki.

## Results

### Sociodemographic characteristics of participants

A total of 327 caregivers were recruited. The majority were women (85.0%), with a significant portion aged between 41 and 50 years (29.4%). Most caregivers were married (58.7%) and had attained a secondary education level (47.7%). In terms of employment, the majority were self-employed (45.3%), with a monthly income typically between 50,000 and 100,000 CFA francs (28.1%) (Table 1).

**Table 1.** Sociodemographic characteristics of study participants, Yaoundé, 2025, Cameroon (n = 327)

| Characteristic | Count (n) | Frequency (%) |
| --- | --- | --- |
| Age |  |  |
| 21 – 30 | 55 | 16.8 |
| 31 – 40 | 73 | 22.3 |
| 41 – 50 | 96 | 29.4 |
| 51 – 60 | 81 | 24.8 |
| 61 – 70 | 22 | 6.7 |
| **Sex** |  |  |
| Female | 278 | 85.0 |
| Male | 49 | 15.0 |
| **Study level** |  |  |
| No formal education | 33 | 10.1 |
| Primary | 85 | 26.0 |
| Secondary | 156 | 47.7 |
| Tertiary | 53 | 16.2 |
| **Marital status** |  |  |
| Single | 99 | 30.3 |
| Divorced | 6 | 1.8 |
| Married | 192 | 58.7 |
| Widow(er) | 30 | 9.2 |
| **Occupation** |  |  |
| Unemployed | 98 | 30.0 |
| Private sector | 54 | 16.5 |
| Public sector | 27 | 8.3 |
| Self-employed | 148 | 45.3 |
| **Income (XAF)** |  |  |
| None | 98 | 30.0 |
| □ 50000 | 69 | 21.1 |
| 50000-100000 | 92 | 28.1 |
| > 100000 | 68 | 20.8 |

XAF: Central Africa CFA Franc

### Exposure to body fluids and vaccination coverage

A low proportion of 10.40% of caregivers had received the full hepatitis B vaccine, 6.12% received the full COVID-19 vaccine, and 3.98% received full cholera vaccine (Table 2). In terms of exposure, 188/327 (57%) participants reported having been exposed to blood and biological fluids during their stay at the hospital. They were most frequently exposed to urine (87%), expectoration (38%), and vomiting (21%) (Fig. 1).

**Table 2.**
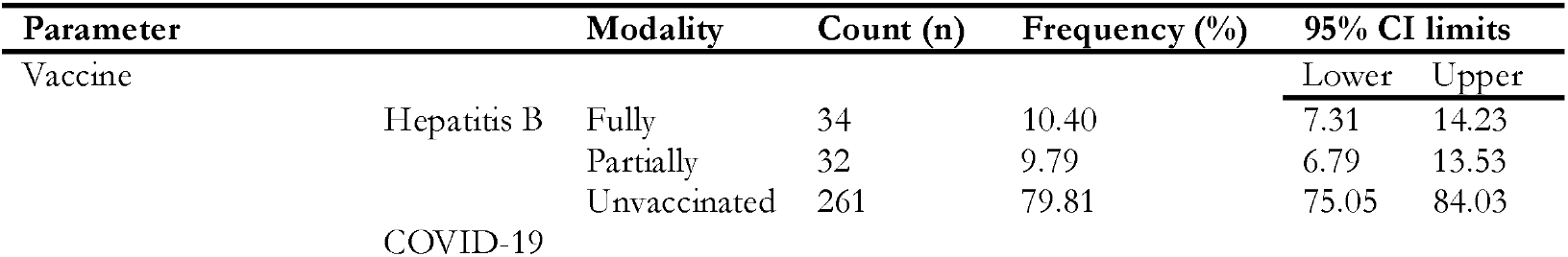
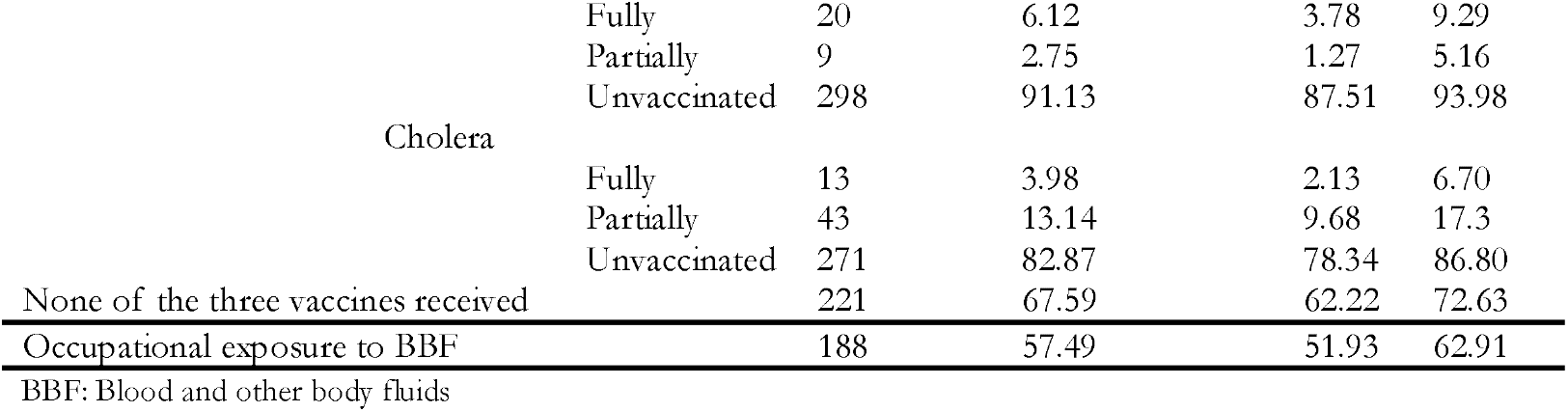
Vaccination status of caregivers in the hospital setting, Yaoundé, 2025, Cameroon (n = 327)

**Fig. 1.**
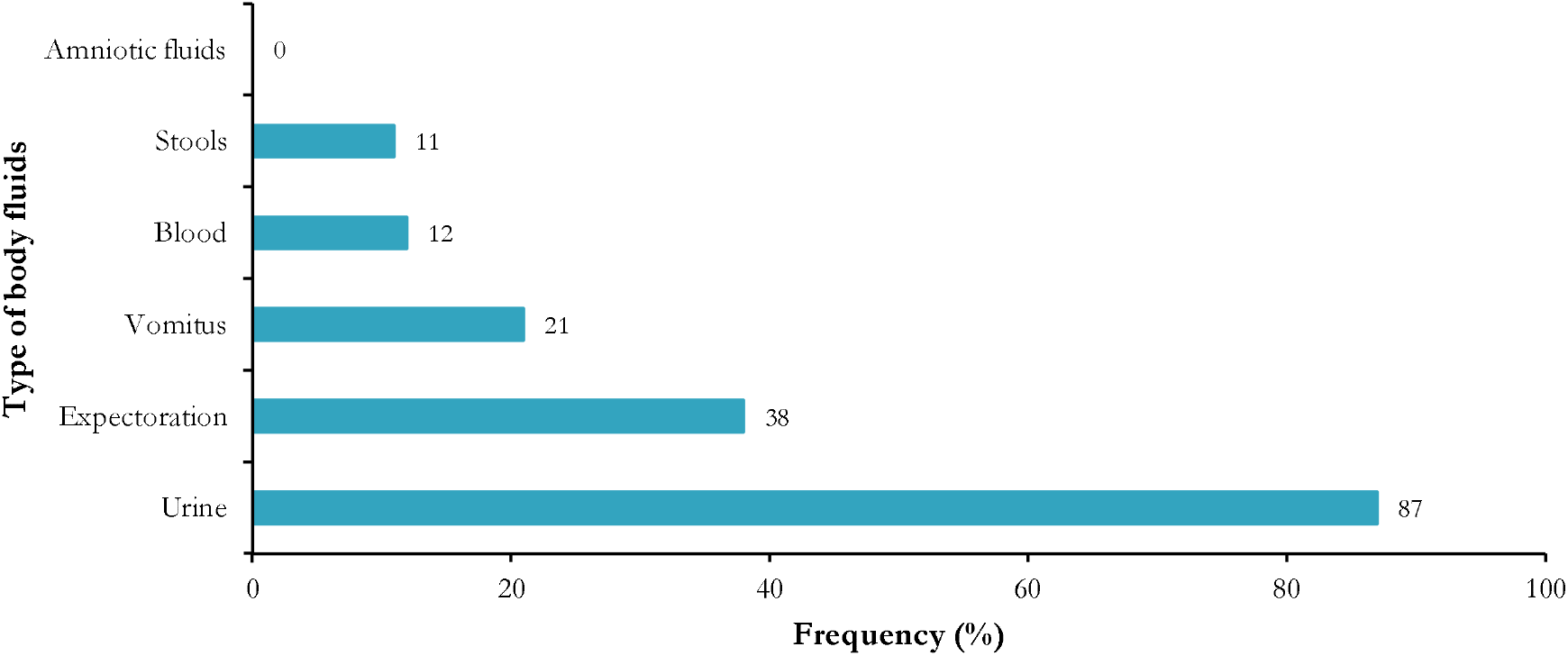
Reported source of hospital exposure to blood and biological fluids experienced by patients’ caregivers in Yaoundé health facilities, 2025, Cameroon

In All, 225/327 (69%) participants reported having previously being caregivers et the hospital. Among them, 88 / 225 (39.11%) stated that they had fallen ill during their experience as caregivers. The most frequently mentioned illnesses were malaria (72%), colds (42%), coughs (36%), and back pain (13%) (Fig. 2).

**Fig. 2.**
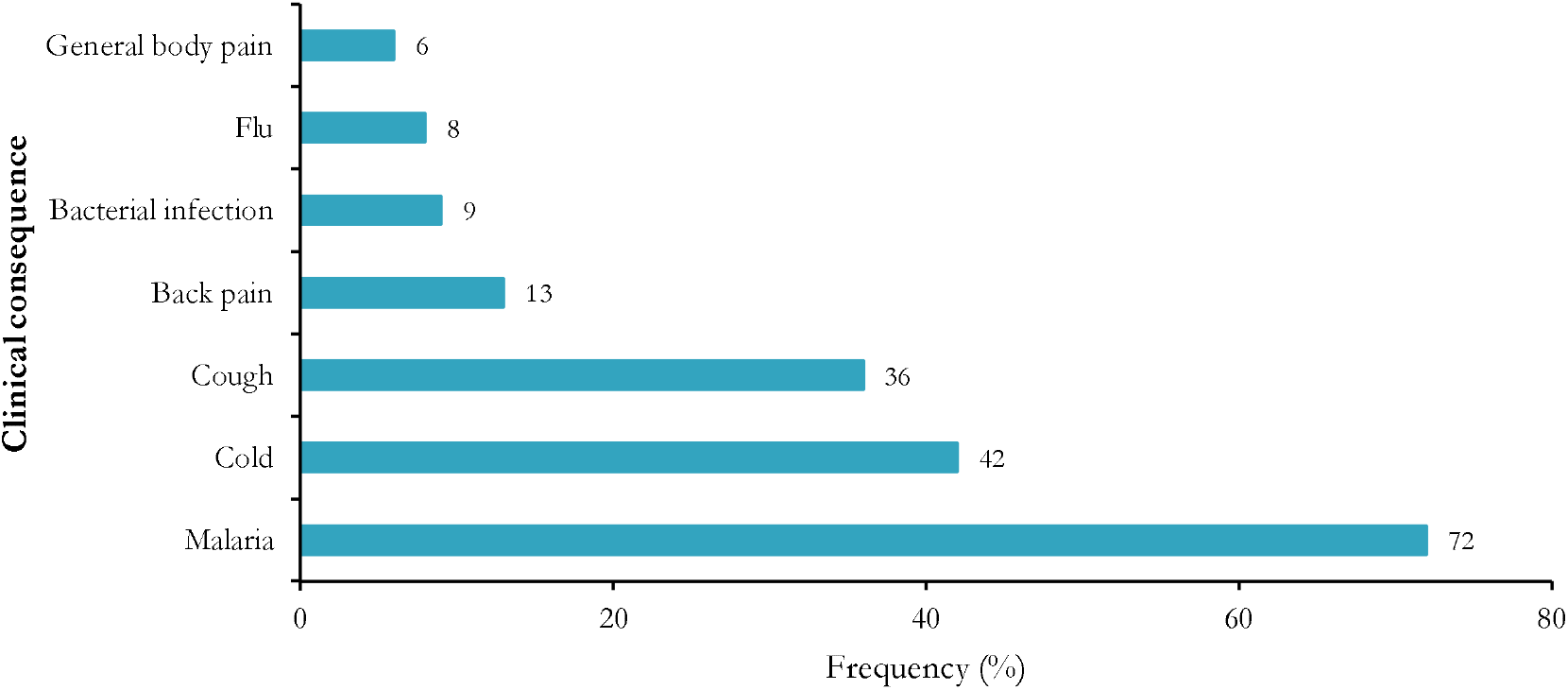
Reported illness experienced after caregiving their patient in Yaoundé health facilities, 2025, Cameroon

### Predictor of preventive vaccination

#### COVID-19 vaccination

Multivariate analysis showed a significant association between gender, marital status, education level, occupation, and COVID-19 vaccine uptake.

Gender: Female caregivers were significantly six times less likely to be fully vaccinated than their male counterparts (AOR = 6.27; 95% CI: 1.42-27.2; *p* = 0.013).

Marital Status: Unmarried caregivers were significantly eight times less likely to be fully vaccinated compared to married caregivers (AOR = 7.90; 95% CI: 2.22-36.3; *p* = 0.003).

Education Level: compared to those with no formal education, caregivers with a higher education level were significantly twenty-nine times less likely to have received a full course of a COVID-19 vaccine (AOR = 29.7; 95% CI: 1.83-65.9; *p* = 0.021) (Table 3).

**Table 3.**
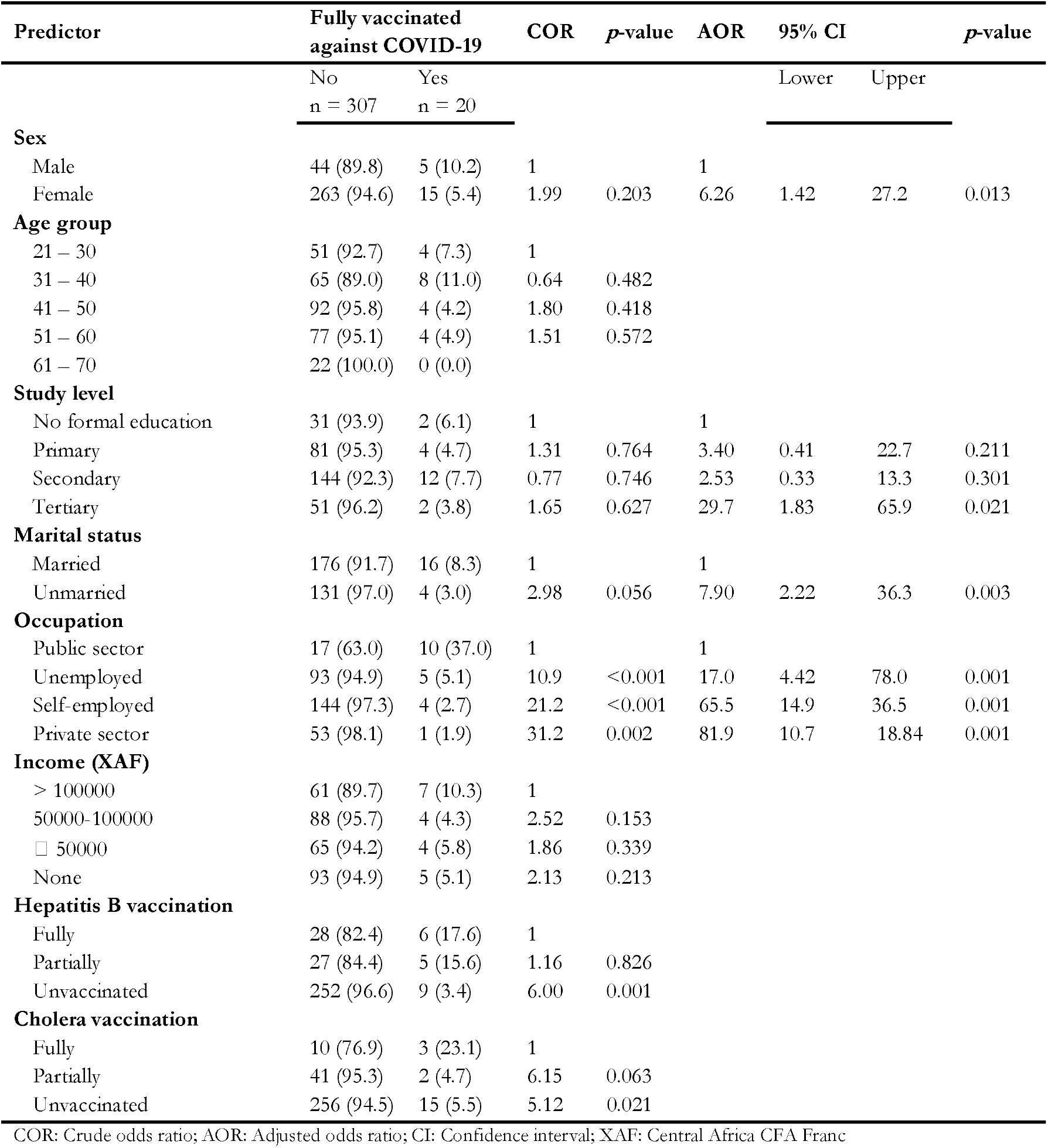
Simple and multiple binary logistic regression of factors associated with COVID-19 full vaccination among caregivers in hospital settings, Yaoundé, 2025, Cameroon (n = 327)

| Predictor | Fully vaccinated<br>against COVID-19 |  | COR | *p*-value | AOR | 95% CI |  | *p*-value |
| --- | --- | --- | --- | --- | --- | --- | --- | --- |
|  | No<br>n = 307 | Yes<br>n = 20 |  |  |  | Lower | Upper |  |
| **Sex** |  |  |  |  |  |  |  |  |
| Male | 44 (89.8) | 5 (10.2) | 1 |  | 1 |  |  |  |
| Female | 263 (94.6) | 15 (5.4) | 1.99 | 0.203 | 6.26 | 1.42 | 27.2 | 0.013 |
| **Age group** |  |  |  |  |  |  |  |  |
| 21 – 30 | 51 (92.7) | 4 (7.3) | 1 |  |  |  |  |  |
| 31 – 40 | 65 (89.0) | 8 (11.0) | 0.64 | 0.482 |  |  |  |  |
| 41 – 50 | 92 (95.8) | 4 (4.2) | 1.80 | 0.418 |  |  |  |  |
| 51 – 60 | 77 (95.1) | 4 (4.9) | 1.51 | 0.572 |  |  |  |  |
| 61 – 70 | 22 (100.0) | 0 (0.0) |  |  |  |  |  |  |
| **Study level** |  |  |  |  |  |  |  |  |
| No formal education | 31 (93.9) | 2 (6.1) | 1 |  | 1 |  |  |  |
| Primary | 81 (95.3) | 4 (4.7) | 1.31 | 0.764 | 3.40 | 0.41 | 22.7 | 0.211 |
| Secondary | 144 (92.3) | 12 (7.7) | 0.77 | 0.746 | 2.53 | 0.33 | 13.3 | 0.301 |
| Tertiary | 51 (96.2) | 2 (3.8) | 1.65 | 0.627 | 29.7 | 1.83 | 65.9 | 0.021 |
| **Marital status** |  |  |  |  |  |  |  |  |
| Married | 176 (91.7) | 16 (8.3) | 1 |  | 1 |  |  |  |
| Unmarried | 131 (97.0) | 4 (3.0) | 2.98 | 0.056 | 7.90 | 2.22 | 36.3 | 0.003 |
| **Occupation** |  |  |  |  |  |  |  |  |
| Public sector | 17 (63.0) | 10 (37.0) | 1 |  | 1 |  |  |  |
| Unemployed | 93 (94.9) | 5 (5.1) | 10.9 | <0.001 | 17.0 | 4.42 | 78.0 | 0.001 |
| Self-employed | 144 (97.3) | 4 (2.7) | 21.2 | <0.001 | 65.5 | 14.9 | 36.5 | 0.001 |
| Private sector | 53 (98.1) | 1 (1.9) | 31.2 | 0.002 | 81.9 | 10.7 | 18.84 | 0.001 |
| **Income (XAF)** |  |  |  |  |  |  |  |  |
| > 100000 | 61 (89.7) | 7 (10.3) | 1 |  |  |  |  |  |
| 50000-100000 | 88 (95.7) | 4 (4.3) | 2.52 | 0.153 |  |  |  |  |
| □ 50000 | 65 (94.2) | 4 (5.8) | 1.86 | 0.339 |  |  |  |  |
| None | 93 (94.9) | 5 (5.1) | 2.13 | 0.213 |  |  |  |  |
| **Hepatitis B vaccination** |  |  |  |  |  |  |  |  |
| Fully | 28 (82.4) | 6 (17.6) | 1 |  |  |  |  |  |
| Partially | 27 (84.4) | 5 (15.6) | 1.16 | 0.826 |  |  |  |  |
| Unvaccinated | 252 (96.6) | 9 (3.4) | 6.00 | 0.001 |  |  |  |  |
| **Cholera vaccination** |  |  |  |  |  |  |  |  |
| Fully | 10 (76.9) | 3 (23.1) | 1 |  |  |  |  |  |
| Partially | 41 (95.3) | 2 (4.7) | 6.15 | 0.063 |  |  |  |  |
| Unvaccinated | 256 (94.5) | 15 (5.5) | 5.12 | 0.021 |  |  |  |  |

COR: Crude odds ratio; AOR: Adjusted odds ratio; CI: Confidence interval; XAF: Central Africa CFA Franc

#### Hepatitis B vaccination

Multivariate analysis revealed a significant association between gender, occupation, and cholera vaccination status with full vaccination against hepatitis B.

Gender: Female caregivers were significantly three times less likely to be fully vaccinated against hepatitis B than male counterparts (AOR = 2.98; 95% CI: 1.16-7.64; *p* = 0.022).

Occupation: Unemployed caregivers were significantly six times less likely to be fully vaccinated for hepatitis B than public sector workers (AOR = 6.43; 95% CI: 1.67-26.1; *p* = 0.007).

Cholera vaccination status: Caregivers who had not received any cholera vaccine were significantly eight times less likely to be fully vaccinated for hepatitis B (AOR = 8.20; 95% CI: 1.76-34.7; *p* = 0.005) (Table 4).

**Table 4.** Simple and multiple binary logistic regression of factors associated with hepatitis B full vaccination among caregivers in hospital sittings, Yaoundé, 2025, Cameroon (n = 327)

| Predictor | Fully vaccinated<br>against hepatitis B |  | COR | p-value | AOR | 95% CI |  | p-value |
| --- | --- | --- | --- | --- | --- | --- | --- | --- |
|  | No<br>n = 283 | Yes<br>n = 34 |  |  |  | Lower | Upper |  |
| **Sex** |  |  |  |  |  |  |  |  |
| Male | 38 (77.6) | 11 (22.4) | 1 |  | 1 |  |  |  |
| Female | 255 (91.7) | 23 (8.3) | 3.21 | 0.004 | 2.98 | 1.16 | 7.64 | 0.022 |
| **Age group** |  |  |  |  |  |  |  |  |
| 21 – 30 | 50 (90.9) | 5 (9.1) | 1 |  | 1 |  |  |  |
| 31 – 40 | 58 (79.5) | 15 (20.5) | 0.39 | 0.085 | 0.36 | 0.09 | 1.24 | 0.122 |
| 41 – 50 | 89 (92.7) | 7 (7.3) | 1.27 | 0.695 | 0.93 | 0.21 | 3.80 | 0.926 |
| 51 – 60 | 79 (97.5) | 2 (2.5) | 3.95 | 0.109 | 2.51 | 0.40 | 21.0 | 0.343 |
| 61 – 70 | 17 (77.3) | 5 (22.7) | 0.34 | 0.119 | 0.27 | 0.05 | 1.37 | 0.110 |
| **Study level** |  |  |  |  |  |  |  |  |
| Tertiary | 42 (79.2) | 11 (20.8) | 1 |  |  |  |  |  |
| Secondary | 141 (90.4) | 15 (9.6) | 2.62 | 0.038 |  |  |  |  |
| Primary | 80 (94.1) | 5 (5.9) | 4.19 | 0.012 |  |  |  |  |
| No formal education | 30 (90.9) | 3 (9.1) | 2.62 | 0.165 |  |  |  |  |
| **Marital status** |  |  |  |  |  |  |  |  |
| Married | 118 (87.4) | 17 (12.6) | 1 |  |  |  |  |  |
| Unmarried | 175 (91.1) | 17 (8.9) | 1.48 | 0.278 |  |  |  |  |
| **Occupation** |  |  |  |  |  |  |  |  |
| Public sector | 20 (74.1) | 7 (25.9) | 1 |  | 1 |  |  |  |
| Private sector | 44 (81.5) | 10 (18.5) | 1.54 | 0.442 | 2.05 | 0.59 | 7.06 | 0.252 |
| Self-employed | 137 (92.6) | 11 (7.4) | 4.36 | 0.006 | 3.93 | 1.18 | 12.6 | 0.022 |
| Unemployed | 92 (93.9) | 6 (6.1) | 5.37 | 0.006 | 6.43 | 1.67 | 26.1 | 0.007 |
| **Income (in XAF)** |  |  |  |  |  |  |  |  |
| > 100000 | 54 (79.4) | 14 (20.6) | 1 |  |  |  |  |  |
| 50000-100000 | 85 (92.4) | 7 (7.6) | 3.15 | 0.020 |  |  |  |  |
| □ 50000 | 62 (89.9) | 7 (10.1) | 2.30 | 0.096 |  |  |  |  |
| None | 92 (93.9) | 6 (6.1) | 3.98 | 0.008 |  |  |  |  |
| **Cholera vaccination** |  |  |  |  |  |  |  |  |
| Fully | 9 (69.2) | 4 (30.8) | 1 |  | 1 |  |  |  |
| Partially | 35 (81.4) | 8 (18.6) | 1.94 | 0.354 | 3.13 | 0.59 | 16.1 | 0.170 |
| Unvaccinated | 249 (91.9) | 22 (8.1) | 5.03 | 0.012 | 8.20 | 1.76 | 34.7 | 0.005 |

COR: Crude odds ratio; AOR: Adjusted odds ratio; CI: Confidence interval; XAF: Central Africa CFA Franc

#### Cholera vaccination

The female gender was significantly three times less likely to be fully vaccinated against cholera than male (Table 5).

**Table 5.** Simple and multiple binary logistic regression of factors associated with cholera vaccination among caregivers in hospital settings, Yaoundé, 2025, Cameroon (n = 327).

| Predictor | Fully vaccinated<br>against Cholera |  | COR | p-value | AOR | 95% CI |  | p-value |
| --- | --- | --- | --- | --- | --- | --- | --- | --- |
|  | No<br>n = 314 | Yes<br>n = 12 |  |  |  | Lower | Upper |  |
| **Sex** |  |  |  |  |  |  |  |  |
| Male | 47 (95.9) | 2 (4.1) |  |  |  |  |  |  |
| Female | 267 (96.0) | 11 (4.0) | 1.03 | 0.967 | 3.36 | 1.07 | 12.6 | 0.048 |
| **Age group** |  |  |  |  |  |  |  |  |
| 21 – 30 | 50 (90.9) | 5 (9.1) |  |  |  |  |  |  |
| 31 – 40 | 72 (98.6) | 1 (1.4) | 7.20 | 0.076 |  |  |  |  |
| 41 – 50 | 92 (95.8) | 4 (4.2) | 2.30 | 0.230 |  |  |  |  |
| 51 – 60 | 79 (97.5) | 2 (2.5) | 3.95 | 0.109 |  |  |  |  |
| 61 – 70 | 21 (95.5) | 1 (4.5) | 2.10 | 0.510 |  |  |  |  |
| **Study level** |  |  |  |  |  |  |  |  |
| Tertiary | 50 (94.3) | 3 (5.7) |  |  |  |  |  |  |
| Secondary | 150 (96.2) | 6 (3.8) | 1.50 | 0.576 |  |  |  |  |
| Primary | 82 (96.5) | 3 (3.5) | 1.64 | 0.554 |  |  |  |  |
| No formal education | 32 (97.0) | 1 (3.0) | 1.92 | 0.579 |  |  |  |  |
| **Marital status** |  |  |  |  |  |  |  |  |
| Married | 126 (93.3) | 9 (6.7) |  |  |  |  |  |  |
| Unmarried | 188 (97.9) | 4 (2.1) | 3.36 | 0.048 |  |  |  |  |
| **Occupation** |  |  |  |  |  |  |  |  |
| Unemployed | 92 (93.9) | 6 (6.1) |  |  |  |  |  |  |
| Self-employed | 144 (97.3) | 4 (2.7) | 2.35 | 0.195 |  |  |  |  |
| Private sector | 52 (96.3) | 2 (3.7) | 1.70 | 0.527 |  |  |  |  |
| Public sector | 26 (96.3) | 1 (3.7) | 1.70 | 0.632 |  |  |  |  |
| **Income (in XAF)** |  |  |  |  |  |  |  |  |
| > 100000 | 65 (95.6) | 3 (4.4) |  |  |  |  |  |  |
| 50000-100000 | 91 (98.9) | 1 (1.1) | 4.20 | 0.218 |  |  |  |  |
| □ 50000 | 66 (95.7) | 3 (4.3) | 1.02 | 0.985 |  |  |  |  |
| None | 92 (93.9) | 6 (6.1) | 0.71 | 0.634 |  |  |  |  |

COR: Crude odds ratio; AOR: Adjusted odds ratio; CI: Confidence interval; XAF: Central Africa CFA Franc
COR: Crude odds ratio; AOR: Adjusted odds ratio; CI: Confidence interval; XAF: Central Africa CFA Franc

#### Risk of exposure to blood and biological fluids among caregivers

More than half of the respondents who reported exposure to body fluids have not received any vaccine course of against hepatitis B (n = 168/327; 51,4%), cholera (n = 178/327; 54.4%) or COVID-19 (n = 173/327; 54.1%).

Multivariate analysis showed a significant association between hospital exposure to blood and biological fluids and several caregiver characteristics:

Prior hospital tasks: Caregivers who had previously performed tasks in the hospital were 17 times more likely to be exposed to blood and biological fluids (AOR = 17.1; 95% CI: 8.36–38.3; *p* < 0.001).

Past caregiving experience: Those with prior caregiving experience were 3 times more likely to be exposed (AOR = 2.98; 95% CI: 1.60–5.57; *p* = 0.001).

Marital status: Married caregivers (AOR = 2.28; 95% CI: 1.24-4.21; *p* = 0.008) were more likely to be exposed to biological fluids during caregiving.

Sleeping condition: caregivers who reported difficulty of sleeping were two to three times more likely to get exposed to biological fluids (AOR = 2.80; 95% CI:1.01-7.81; *p* = 0.047 and AOR = 3.28; 95% CI: 1.21-8.97; *p* = 0.019). (Table 6)

**Table 6.** Simple and multivariate regression of caregivers’ exposure risk to blood and biological fluids in Yaoundé hospital settings, 2025, Cameroon (n = 327)

| Risk factor | Exposed to BBF |  | COR | *p*-value | AOR | 95% CI |  | *p*-value |
| --- | --- | --- | --- | --- | --- | --- | --- | --- |
|  | Yes<br>n = 188 | No<br>n = 139 |  |  |  | Lower | Upper |  |
| **Sex** |  |  |  |  |  |  |  |  |
| Male | 20 (40.8) | 29 (59.2) | 1 |  |  |  |  |  |
| Female | 168 (60.4) | 110 (39.6) | 2.21 | 0.012 |  |  |  |  |
| **Age group** |  |  |  |  |  |  |  |  |
| 21 – 30 | 18 (32.7) | 37 (67.3) | 1 |  |  |  |  |  |
| 31 – 40 | 35 (47.9) | 38 (52.1) | 1.89 | 0.085 |  |  |  |  |
| 41 – 50 | 58 (60.4) | 38 (39.6) | 3.14 | 0.001 |  |  |  |  |
| 51 – 60 | 59 (72.8) | 22 (27.2) | 5.51 | <0.001 |  |  |  |  |
| 61 – 70 | 18 (81.8) | 4 (18.2) | 9.25 | <0.001 |  |  |  |  |
| **Study level** |  |  |  |  |  |  |  |  |
| Tertiary | 22 (41.5) | 31 (58.5) | 1 |  |  |  |  |  |
| Secondary | 91 (58.3) | 65 (41.7) | 1.97 | 0.035 |  |  |  |  |
| Primary | 52 (61.2) | 33 (38.8) | 2.22 | 0.025 |  |  |  |  |
| No formal education | 23 (69.7) | 10 (30.3) | 3.24 | 0.012 |  |  |  |  |
| **Marital status** |  |  |  |  |  |  |  |  |
| Single | 40 (40.4) | 59 (59.6) | 1 |  | 1 |  |  |  |
| Divorced | 2 (33.3) | 4 (66.7) | 0.74 | 0.732 | 0.28 | 0.04 | 1.83 | 0.188 |
| Married | 119 (62.0) | 73 (38.0) | 2.40 | 0.001 | 2.28 | 1.24 | 4.21 | 0.008 |
| Widow(er) | 27 (90.0) | 3 (10.0) | 13.3 | 0.001 | 14.0 | 3.51 | 75.3 | <0.001 |
| **Occupation** |  |  |  |  |  |  |  |  |
| Public sector | 7 (25.9) | 20 (74.1) | 1 |  |  |  |  |  |
| Private sector | 29 (53.7) | 25 (46.3) | 3.31 | 0.020 |  |  |  |  |
| Self-employed | 97 (65.5) | 51 (34.5) | 5.43 | <0.001 |  |  |  |  |
| Unemployed | 55 (56.1) | 43 (43.9) | 3.65 | 0.007 |  |  |  |  |
| **Income (In XAF)** |  |  |  |  |  |  |  |  |
| None | 55 (56.1) | 43 (43.9) | 1 |  |  |  |  |  |
| □ 50000 | 39 (56.5) | 30 (43.5) | 1.02 | 0.959 |  |  |  |  |
| 50000-100000 | 54 (58.7) | 38 (41.3) | 1.11 | 0.720 |  |  |  |  |
| > 100000 | 40 (58.8) | 28 (41.2) | 1.12 | 0.729 |  |  |  |  |
| **Caregiving period (in days)** |  |  |  |  |  |  |  |  |
| 2-3 | 33 (37.9) | 54 (62.1) | 1 |  |  |  |  |  |
| 4-7 | 98 (58.7) | 69 (41.3) | 2.32 | 0.002 |  |  |  |  |
| 8+ | 57 (78.1) | 16 (21.9) | 5.83 | <0.001 |  |  |  |  |
| **Had trouble with a HCW** |  |  |  |  |  |  |  |  |
| No | 126 (54.5) | 105 (45.5) | 1 |  |  |  |  |  |
| Yes | 62 (64.6) | 34 (35.4) | 1.52 |  |  |  |  |  |
| **Hepatitis B vaccination** |  |  |  |  |  |  |  |  |
| Fully | 20 (58.8) | 14 (41.2) | 1 |  |  |  |  |  |
| Partially | 17 (53.1) | 15 (46.9) | 0.79 | 0.641 |  |  |  |  |
| Unvaccinated | 151 (57.9) | 110 (42.1) | 0.96 | 0.914 |  |  |  |  |
| **COVID-19 vaccination** |  |  |  |  |  |  |  |  |
| Fully | 11 (55.0) | 9 (45.0) | 1 |  |  |  |  |  |
| Partially | 3 (33.3) | 6 (66.7) | 0.41 | 0.286 |  |  |  |  |
| Unvaccinated | 174 (58.4) | 124 (41.6) | 1.15 | 0.766 |  |  |  |  |
| **Cholera vaccination** |  |  |  |  |  |  |  |  |
| Fully | 10 (76.9) | 3 (23.1) | 1 |  |  |  |  |  |
| Partially | 26 (60.5) | 17 (39.5) | 0.46 | 0.285 |  |  |  |  |
| Unvaccinated | 152 (56.1) | 119 (43.9) | 0.38 | 0.152 |  |  |  |  |
| **Performed task during the hospital stay** |  |  |  |  |  |  |  |  |
| No | 12 (14.0) | 74 (86.0) | 1 |  | 1 |  |  |  |
| Yes | 176 (73.0) | 65 (27.0) | 16.7 | <0.001 | 17.1 | 8.36 | 38.3 | <0.001 |
| **Have been caregiver before** |  |  |  |  |  |  |  |  |
| No | 33 (32.4) | 69 (67.6) | 1 |  | 1 |  |  |  |
| Yes | 155 (68.9) | 70 (31.1) | 4.63 | <0.001 | 2.98 | 1.60 | 5.57 | 0.001 |
| **Feel easy to sleep during hospital stay** |  |  |  |  |  |  |  |  |
| Yes | 11 (40.7) | 16 (59.3) | 1 |  | 1 |  |  |  |
| Sometime | 78 (59.5) | 53 (40.5) | 2.14 | 0.077 | 2.80 | 1.01 | 7.81 | 0.047 |
| Never | 99 (58.6) | 70 (41.4) | 2.06 | 0.087 | 3.28 | 1.21 | 8.97 | 0.019 |

BBF: Blood and other body fluids; COR: Crude odds ratio; AOR: Adjusted odds ratio; CI: Confidence interval; XAF: Central Africa CFA Franc; HCW: Healthcare worker

## Discussion

This study aimed to provide a descriptive overview and identify the determinants of exposure to blood and body fluids (BBF) among caregivers, as well as to assess their vaccination status. The results reveal that caregivers are highly exposed to these fluids in the hospital setting, despite very low vaccination coverage.

The majority of participants were female and over 41 years old. Similar results were found in a study conducted in Iran, which could be explained by the fact that as women get older, they are more likely to care for their relatives without necessarily considering the risks of exposure [25]. Half of the participants were either unemployed or self-employed and had a monthly income of less than 50,000 CFA francs. Indeed, due to the long hospital stays required, families tend to assign the most available person to stay by the patient’s side [17].

Among participants who had previously been patient attendants, over 25% had contracted an illness during their hospital stay. This concerning figure corroborates the findings of a Cameroonian study documenting the precarious living conditions of these caregivers [17,26]. Due to a lack of suitable accommodation, they are often compelled to sleep on benches, in corridors, or in the open air, which significantly increases their risk of exposure to pathogens. This situation calls for urgent action from policymakers to guarantee the health and well-being of this vulnerable population by creating safe and suitable reception areas within healthcare facilities.

Just under a quarter of the caregivers surveyed had received a complete COVID-19 vaccination schedule, a finding similar to that observed in Congo [27]. This low coverage is mainly explained by vaccine hesitancy, fueled by doubts about the vaccines’ efficacy and safety, a lack of information, and widespread mistrust [28,29]. To promote uptake, vaccination strategies must therefore imperatively target the specific factors underlying this hesitancy within the population [30].

A higher level of education was significantly associated with COVID-19 vaccination. This trend was also observed in a study conducted in the United States [31]. Caregivers with a higher level of education are indeed more likely to adopt health-promoting behaviors, including preventive measures against infectious diseases like COVID-19. Furthermore, they generally benefit from better access to health resources, particularly information on disease prevention [32]. These results underscore the need to strengthen vaccination awareness, specifically targeting less educated populations. To effectively reach these groups, including in the most remote areas, it is essential to use widely accessible communication channels, such as media (television, radio), social networks, and health facilities.

Regarding hepatitis B, only 10.40% of caregivers had received a complete vaccination. This insufficient vaccination coverage worsens the burden of this preventable disease [33]. This result is lower than that observed in another Cameroonian study, a discrepancy that could be explained by differences in the study populations [7].

Studies have identified several barriers that could explain this low vaccination coverage, including the high cost of the vaccine, lack of time to get vaccinated, neglect, and vaccine unavailability [7,34]. To reduce the incidence of infection, the policy of free vaccination for newborns of infected mothers must be reinforced by intensive behavior change campaigns aimed at promoting preventive behaviors within the population [35].

Unemployed caregivers and those not vaccinated against cholera were respectively six and eight times more likely to be vaccinated against hepatitis B. This trend, which contrasts with data from an Ethiopian study, could reveal a unique opportunity to target these groups in future awareness and vaccination strategies [36].

Less than 5% of caregivers were fully vaccinated against cholera, with a lack of awareness of the vaccine’s existence identified as a major barrier [14]. Consequently, it is crucial to intensify awareness campaigns focused on communities and to adjust public health strategies to better meet the specific needs of this population [37].

Over 50% of caregivers were exposed to body fluids, a result similar to that observed in Bangladesh [8]. Most of the exposed caregivers were not fully vaccinated against hepatitis B, Cholera, and COVID-19. This makes them more vulnerable to infectious diseases, especially if an outbreak appears in the study area [38,39].

These findings corroborate results among healthcare workers in both a district and a referral hospital in Yaoundé. Workers responsible for providing primary care were not fully protected against these diseases, a situation worsened by recurrent shortages of personal protective equipment, poor compliance with standard precautions, and inadequate management of exposure cases in most hospital settings [39–43].

Multivariate analysis revealed that caregivers performing hospital tasks had a 17-fold higher risk of exposure to blood and body fluids, while those with prior caregiving experience had a three-fold higher risk. Paradoxically, these essential care actors receive no training in basic precautions, leaving them defenseless against the growing threat of nosocomial infections [5,44].

### Limitations

The main limitation of this study is the risk of recall bias regarding previous exposure, or the difficulty in proving that exposure to the risk was prior to the outcome. The study was also subject to social desirability bias but this was mitigated by asking multiple question to establish the vaccination status of the respondent (type of vaccine, number of doses received and the approximate date of the last dose). In addition, a qualitative research design might be helpful in understanding why help understanding the deep reason behind poor compliance with vaccination and other issues raised in this study.

## Conclusions

Caregivers play an indispensable role in the Cameroonian healthcare system. They perform their tasks without protective measures and are at a significant risk of exposure to blood and other body fluids. Vaccination coverage among these caregivers is very low for diseases such as hepatitis B, COVID-19, and cholera, with less than a quarter being fully vaccinated. Furthermore, their precarious living conditions are so dire that a significant proportion have already contracted illnesses during their hospital stays, primarily malaria, respiratory infections (colds, coughs), and back pain. Given this critical situation, it is urgent for policymakers to integrate informal caregivers as key players in the health system. This requires implementing appropriate policies and providing them with the necessary tools to reduce their exposure to infectious risks. These actions should include creating safe reception areas, providing training on risk protection within hospitals, and intensifying targeted awareness campaigns to improve prevention and vaccination.

## Data Availability

All data generated or analyzed during this study are included in this published article.

## Abbreviations

*a*OR: Adjusted Odds Ration
*CI*: Confidence Interval
*c*OR: Crude Odds Ratio
*COVID-19*: New Coronavirus Disease

## Declaration

### Authors’ contribution

Conception, drafting of the study protocol, data collection, analysis, and interpretation: R.T.; Drafting and editing of manuscript: R.T. and F.Z.L.C.; Critical revision of the manuscript: R.T., A.E.M., G.A.M., G.R.E. and F.Z.L.C.; Conception, design and supervision of research protocol and implementation, data analysis plan, data analysis, interpretation, revision, and editing: F.Z.L.C.; Final validation of the manuscript: All authors.

### Consent for publication

Not applicable.

### Availability of data and materials

All data generated or analyzed during this study are included in this published article.

## Acknowledgements

Our gratitude goes to the manager of these health facilities who gave an authorization to conduct the study.

## References

1. Oleribe OO, Momoh J, Uzochukwu BS, Mbofana F, Adebiyi A, Barbera T, et al. Identifying Key Challenges Facing Healthcare Systems In Africa And Potential Solutions. Int J Gen Med. 2019 Nov 6; 12:395–403.

2. Cheuyem FZL, Amani A, Ajong BN, Boukeng LBK, Mouangue C, Tsafack MGM, et al. Humanization of Care: A Geospatial Analysis of Key Indicators of Quality and Safety of Health Care and Service in the Centre Region, Cameroon. medRxiv; 2024. doi:10.1101/2024.11.11.24317125v1.

3. Willcox ML, Peersman W, Daou P, Diakité C, Bajunirwe F, Mubangizi V, et al. Human resources for primary health care in sub-Saharan Africa: progress or stagnation? Hum Resour Health. 2015;13(1):76.

4. Ze A, Tsele FPN. Sustainable Development Goal No. 3 of 2030 Agenda: An Evaluation of Universal Health Coverage Level Achievement in Cameroon. Cent Afr J Public Health. 2025;11(4):154–64.

5. Hailu GN, Gebru HB, Hagos GG, Weldemariam AH, Tadesse DB, Mebrahtom G. The role of family caregivers in supporting older adults in Africa: systematic review. BMC Geriatr. 2025;25(1):491.

6. Westermann C, Peters C, Lisiak B, Lamberti M, Nienhaus A. The prevalence of hepatitis C among healthcare workers: a systematic review and meta-analysis. Occup Environ Med. 2015;72(12):880–8.

7. Cheuyem FZL, Lyonga EE, Kamga HG, Mbopi-Keou FX, Takougang I. Needlestick and Sharp Injuries and Hepatitis B Vaccination among Healthcare Workers: A Cross-Sectional Study in Six District Hospitals in Yaounde (Cameroon). J Community Med Public Health. 2023;7(3):1–9.

8. Islam MS, Luby SP, Sultana R, Rimi NA, Zaman RU, Uddin M, et al. Family caregivers in public tertiary care hospitals in Bangladesh: Risks and opportunities for infection control. Am J Infect Control. 2014;42(3):305–10.

9. Besombes C, Njouom R, Paireau J, Lachenal G, Texier G, Tejiokem M, et al. The epidemiology of hepatitis delta virus infection in Cameroon. 2020; https://gut.bmj.com/content/69/7/1294.abstract. Accessed: 2025 Aug 28.

10. Mboringong AB, Ngomtcho SCH, Ndip Ndip R, Linda EE, Bertand DL, Patricia M, et al. Trends of cholera epidemics and associated mortality factors in Cameroon: 2018–2023: a cross-sectional study. BMC Public Health. 2025;25(1):1816.

11. CAMEROUN Rapport de situation COVID-19 N°212 fr. CCOUSP. https://www.ccousp.cm/download/cameroun-rapport-de-situation-covid-19-n212-fr/. Accessed: 2025 Aug 28.

12. Amani A, Njoh AA, Mouangue C, Zobel CLF, Mossus T. Vaccination Coverage and Safety in Cameroon; Descriptive Assessment of COVID-19 Infection in Vaccinated Individuals. Health Sci Dis. 2022 ;23(8).1–8.

13. Lutte contre l’hépatite B□: le Cameroun choisit d’intervenir dès la naissance. https://www.gavi.org/fr/vaccineswork/lutte-contre-hepatite-b-cameroun-choisit-intervenir-des-naissance. Accessed: 2025 Aug 29.

14. Amani A, Ngo Bama S, Dia M, Nguefack Lekelem S, Linjouom A, Mossi Makembe H, et al. Challenges, best practices, and lessons learned from oral cholera mass vaccination campaign in urban Cameroon during the COVID-19 era. Vaccine. 2022;40(47):6873–9.

15. Cheuyem FZL, Amani A, Nkodo ICA, Boukeng LBK, Edzamba MF, Nouko A, et al. COVID-19 vaccine acceptance and hesitancy in Cameroon: a systematic review and meta-analysis. BMC Public Health. 2025 Mar 17;25(1):1035.

16. Edwige Omona Guissana, Florence Kissougle Nkongo, Metomb Franck Steve, Fabrice Zobel Lekeumo Cheuyem, Ariane Nouko, *et.al*. Oral Cholera Vaccine Acceptability in the Biyem-Assi Health District: A Cross-Sectional Analytical Study in Yaoundé-Cameroon. Am J Biomed Sci & Res. 2024 24(6). doi: 10.34297/AJBSR.2024.24.003264.

17. CSSAC. Les profanes «□professionnels□» de santé□: Le garde malade au coeur de l’organisation du système de santé au Cameroun. Mballa Elanga Edmond VII-Université de Douala-Dpt. https://mballaelanga.canalblog.com/archives/2012/09/05/25052056.html. Accessed: 2025 Aug 1.

18. Cheuyem FZL, Asahngwa CT, Dabou S, Ajong BN, Nloga GS, Goupeyou-Youmsi J, et al. Healthcare coverage and associated factors in Cameroon: analyses from a national survey . medRxiv; 2025. doi: 10.1101/2025.06.22.25330087.

19. Cameroun: Profil Urbain de Yaoundé | UN-Habitat. https://unhabitat.org/cameroun-profil-urbain-de-yaounde-2. Accessed: 2025 Jul 28.

20. Les grands hôpitaux publics. Osidimbea - La Mémoire du Cameroun. Encyclopédie de l’histoire des organisations. [Accessed: 2025 Jul 28]. http://mystory-medical.jimdofree.com/etablissements-cat-1-2/

21. Reports | DHIS2. Ministry of Public Health, Cameroon. 2025. https://dhis-minsante-cm.org/dhis-web-reports/index.html#/data-set-report. Accessed: 2025 Aug 3.

22. Egbuchulem KI. The basics of sample size estimation: an editor’s view. Ann Ib Postgrad Med. 2023;21(1):5–10.

23. R Core Team. R: A Language and Environment for Statistical Computing. R Foundation for Statistical Computing, Vienna, Austria. 2024. https://www.R-project.org/. Accessed: 2024 May 30.

24. Akaike Information Criterion - an overview | ScienceDirect Topics. https://www.sciencedirect.com/topics/pharmacology-toxicology-and-pharmaceutical-science/akaike-information-criterion. Accessed: 2025 Jul 28.

25. Etemadifar S, Heidari M, Jivad N, Masoudi R. Effects of family-centered empowerment intervention on stress, anxiety, and depression among family caregivers of patients with epilepsy. Epilepsy Behav. 2018 Nov 1; 88:106–12.

26. Bleriau Pueugueu I. Repenser la place des “gardes-malades” dans l’équipe de prise en charge en Afrique subsaharienne□: une réflexion sure “l’ethno soins infirmiers.” 2021. http://hdl.handle.net/1866/25726. Accessed: 2025 Jul 29.

27. Guillaume AS, Ndwandwe D, Nyalundja AD, Bugeme PM, Ntaboba AB, Hatu’m VU, et al. Caregivers’ hesitancy and outright refusal toward children’s COVID-19 vaccination in the Democratic Republic of Congo: A community-based cross-sectional study. Hum Vaccines Immunother. 2024;20(1):2422686.

28. Maamor NH, Muhamad NA, Dali NSM, Leman FN, Rosli IA, Shah Tpntb, et al. Prevalence of caregiver hesitancy for vaccinations in children and its associated factors: A systematic review and meta-analysis. PLOS ONE. 2024;19(10): e0302379.

29. Fouda AAB, Kengne VFM, Adiogo D, Manga LJO. Refus et hésitation vis-à-vis de la vaccination anti-COVID-19 à Douala, Cameroun. Pan Afr Med J . 2024 48(1). doi: 10.11604/pamj.2024.48.61.39880.

30. Dinga JN, Njoh AA, Gamua SD, Muki SE, Titanji VPK. Factors Driving COVID-19 Vaccine Hesitancy in Cameroon and Their Implications for Africa: A Comparison of Two Cross-Sectional Studies Conducted 19 Months Apart in 2020 and 2022. Vaccines. 2022 Aug 26;10(9):1401.

31. Khairat S, Zou B, Adler-Milstein J. Factors and reasons associated with low COVID-19 vaccine uptake among highly hesitant communities in the US. Am J Infect Control. 2022;50(3):262–7.

32. Humer E, Jesser A, Plener PL, Probst T, Pieh C. Education level and COVID-19 vaccination willingness in adolescents. Eur Child Adolesc Psychiatry. 2023;32(3):537–9.

33. Pattyn J, Hendrickx G, Vorsters A, Van Damme P. Hepatitis B Vaccines. J Infect Dis. 2021;224(12 Suppl 2): S343–51.

34. Ngum AM, Laure SJ, Tchetnya X, Tambe TA, Ngwayu CN, Wirsiy FS, et al. Vaccination against Hepatitis B among health care workers in the Bamenda Health District: influence of knowledge and attitudes, Cameroon. Pan Afr Med J. 2021; 40:216.

35. StopBlaBlaCam. Hépatite B□: le Cameroun autorise la vaccination systématique des nouveau-nés pour éliminer la transmission mère-enfant. https://www.stopblablacam.com/societe/2202-13808-hepatite-b-le-cameroun-autorise-la-vaccination-systematique-des-nouveau-nes-pour-eliminer-la-transmission-mere-enfant. Accessed: 2025 Aug 8.

36. Haile K, Timerga A, Mose A, Mekonnen Z. Hepatitis B vaccination status and associated factors among students of medicine and health sciences in Wolkite University, Southwest Ethiopia: A cross-sectional study. PLOS ONE. 2021;16(9):e0257621.

37. Omona Guissama E, Kissougle Nkongo F, Metomb FS, Lekeumo Cheuyem FZ, Nouko A, Mbida HS, et al. Oral Cholera Vaccine Acceptability in the Biyem-Assi Health District: A Cross-sectional Analytical study in Yaoundé-Cameroon. Rochester, NY: Social Science Research Network; 2023. https://papers.ssrn.com/abstract=4534818. Accessed: 2025 Aug 29.

38. Cheuyem FZL, Amani A, Achangwa C, Ajong BN, Minkandi CA, Zeh MMMK, et al. COVID-19 vaccine uptake and its determinants in Cameroon: a systematic review and meta-analysis (2021–2024). BMC Infect Dis. 2025;25(1):525.

39. Takougang I, Cheuyem FZL, Lyonga EE, Ndungo JH, Mbopi-Keou FX. Observance of Standard Precautions for Infection Prevention in The Covid-19 Era: A Cross-Sectional Study in Six District Hospitals in Yaounde, Cameroon. Am J Biomed Sci Res. 2023;19(5):590–8.

40. Takougang I, Cheuyem FZL, Ze BRS, Tsamoh FF, Moneboulou HM. Awareness of standard precautions, circumstances of occurrence and management of occupational exposures to body fluids among healthcare workers in a regional level referral hospital (Bertoua, Cameroon). BMC Health Serv Res. 2024;24(1):424.

41. Takougang I, Cheuyem FZL, Ndungo JH, Lyonga EE, Mbopi-Keou FX. Infection Risk Perception, Reporting and Post-Exposure Management of Occupational Injuries among Healthcare Workers in District Hospitals. J Clin Gastroenterol Hepatol. 2023;7(4):1–7.

42. Takougang I, Cheuyem FZL, Changeh BA, Nyonga ND, Moneboulou HM. Accidental Exposure to Body Fluids Among Healthcare Workers in a Referral Hospital in the Security-Challenged Region of South West Cameroon. J Nurs Healthc. 2024;9(2):1–13.

43. Cheuyem FZL, Lyonga EE, Takougang I. Standard precautions perception and practice among health workers in the obstetrics-gynecology department of a referral hospital in Cameroon. BMC Health Serv Res. 2025;25(1):1165.

44. Cheng CC, Fann LY, Chou YC, Liu CC, Hu HY, Chu D. Nosocomial infection and spread of SARS-CoV-2 infection among hospital staff, patients and caregivers. World J Clin Cases. 2022;10(34):12559–65.

